# A Sliding Window Approach to Optimize the Time-varying Parameters of a Spatially-explicit and Stochastic Model of COVID-19

**DOI:** 10.1101/2022.03.21.22272590

**Authors:** Saikanth Ratnavale, Crystal Hepp, Eck Doerry, Joseph R Mihaljevic

## Abstract

The implementation of non-pharmaceutical public health interventions can have simultaneous impacts on pathogen transmission rates as well as host mobility rates. For instance, with SARS-CoV-2, masking can influence host-to-host transmission, while stay-at-home orders can influence mobility. Importantly, variations in transmission rates and mobility patterns can influence pathogen-induced hospitalization rates. This poses a significant challenge for the use of mathematical models of disease dynamics in forecasting the spread of a pathogen; to create accurate forecasts in spatial models of disease spread, we must simultaneously account for time-varying rates of transmission and host movement. In this study, we develop a statistical model-fitting algorithm to estimate dynamic rates of SARS-CoV-2 transmission and host movement from geo-referenced hospitalization data. Using simulated data sets, we then test whether our method can accurately estimate these time-varying rates simultaneously, and how this accuracy is influenced by the spatial population structure. Our model-fitting method relies on a highly parallelized process of grid search and a sliding window technique that allows us to estimate time-varying transmission rates with high accuracy and precision, as well as movement rates with somewhat lower precision. Estimated parameters also had lower precision in more rural data sets, due to lower hospitalization rates (i.e., these areas are less data-rich). This model-fitting routine could easily be generalized to any stochastic, spatially-explicit modeling framework, offering a flexible and efficient method to estimate time-varying parameters from geo-referenced data sets.

## Introduction

In March 2020, the World Health Organization announced that the spread of SARS-CoV-2 virus – the etiological agent of COVID-19 – was officially a pandemic. Various preventive measures such as social distancing, wearing masks, and stay-at-home orders were recommended to contain the outbreak throughout the pandemic, and studies have shown that some non-pharmaceutical public health interventions have been more effective than others (Bertozzi et al. 2020; Drake et al. 2021). At present, vaccinations play a major role in reducing the number of infectious individuals, serious illness, and deaths. Because implementation of prevention measures have varied across space and time, SARS-CoV-2 transmission rates and human mobility rates have changed in local regions throughout the pandemic. This poses a challenge to modeling studies that seek to fit appropriate mathematical models of SARS-CoV-2 transmission to local data for the purposes of hypothesis testing (e.g., evaluating effects of specific interventions) and near-term forecasting (e.g., predicting hospitalization numbers).

As models of disease transmission have become more complex, e.g., spatially-explicit or agent-based models, the challenges of parameterizing such models by fitting them to disease surveillance data have increased as well (Keeling and Rohani 2011). In classic compartmental disease models that are based on ordinary differential equations, such as the Susceptible-Infectious-Removed (SIR) model, only one host population is considered and the pathogen transmission rate is constant over time. However, these assumptions can be relaxed, such that transmission rates can fluctuate over time (e.g., seasonal-forcing), and/or hosts can reside in distinct populations situated within a geographic space that are connected by host movement (i.e., meta-population disease models) (Yang et al. 2021; Yang, Shaff, and Shaman 2021).

Spatially-explicit disease models that also allow for time-varying transmission rates can be computationally expensive and pose a challenge for parameterization (Riley 2007; Riley et al. 2015). For example, various models of SARS-CoV-2 transmission have allowed transmission rates to change over time in response to public health interventions (Bertozzi et al. 2020; Chinazzi et al. 2020). Other models are spatially explicit, including spatial transmission rates (e.g., the probability of transmission between two locations) or have allowed for human mobility rates to influence transmission between locations (Block et al. 2020; Yang, Shaff, and Shaman 2021). A problem with these models, however, is that it is not well known whether they can be fully parameterized: can we simultaneously estimate time-varying movement rates between locations and time-varying transmission rates within populations, where both rates can be influenced by different public health interventions? For instance, lock-downs can limit travel between locations, and masking that can limit the effective contact rate between individuals, and both of these interventions could ultimately impact the number of observed pathogen-related hospitalizations. Moreover, estimating time-varying rates in these models is important not only for providing accurate short-term forecasts, but also for retroactively assessing the quantitative effects of public health interventions or population-level behaviors on observed disease rates.

In this study, we propose statistical methods to efficiently estimate time-varying transmission rates and host movement rates in a stochastic and spatially-explicit model of SARS-CoV-2 transmission. There are many different methodologies for estimating static and dynamic parameters of epidemiological models (e.g., transmission rates or other rates of transitions between compartments, like recovery rates, etc.). Common parameter estimation routines include projection algorithms, gradient algorithms, and algorithms based on least squares (Cantó, Coll, and Sánchez 2017). More recently, Bayesian algorithms and particle filtration methods have been applied to epidemiological problems (Arias et al. 2021; Cazelles, Champagne, and Dureau 2018; King, Nguyen, and Ionides 2015). These estimation algorithms and associated software help epidemiologists understand the dominant drivers of disease transmission, allow for improved forecasting with accurate parameter values, and allow for uncertainty quantification and the propagation of uncertainty into model forecasts. An overview of different parameter estimation methods with their advantages and drawbacks is given by Chou and Voit (2009). Many of these model-fitting strategies, however, do not allow for time-varying parameters of mechanistic models that have explicit spatial structure.

Here we propose using a grid search method, which is an optimization algorithm widely used in machine learning (Liashchynskyi and Liashchynskyi 2019; Shekar and Dagnew 2019), combined with a dynamic programming approach, to efficiently traverse a complicated parameter estimation problem. In grid search, a parameter space is defined, and the algorithm searches many possible combinations of the parameter values to optimize the fit of the model to the data. Dynamic programming is one of the most powerful methods used to solve optimization related problems in mathematics and computer science (Bellman 1954). In this method, a complex problem is simplified by recursively breaking it down into simpler sub-problems such that the larger problem is solved by solving individual sub-problems. The sliding window technique (Braverman 2016; Datar and Motwani 2007) is a fundamental technique of dynamic programming that can be used to tackle optimization problems in which parameters of a model might vary over time. With a sliding window, a sub-problem is defined over a portion of the larger data set, effectively looking at multiple time-slices of the data set. The window is moved over the entire data set while solving sub-problems within each window. This method is frequently applied to solve simple problems including finding local maxima, running averages, sums of sub-arrays, and is also used in more complicated problems involving complex strings of characters (Arasu and Manku 2004; Datar et al. 2002; Faro and Lecroq 2012).

Here we introduce a user-friendly method that combines grid search with the sliding window technique and discuss its time-efficient implementation on a high-performance computing cluster. We then use simulated case studies to the ability of our method simultaneously estimate time-varying rates of transmission and host movement within a stochastic and spatially-explicit model of SARS-CoV-2 and COVID-19 disease dynamics. Results show that our method is able to provide robust parameter estimates, although ability to estimate some parameters precisely is mediated by specific spatial characteristics of the meta-population of interest.

## Methods

### Overview

We start with an epidemiological model that represents the transmission of SARS-CoV-2 and the hospitalization dynamics related to COVID-19 disease, which we have described previously (Mihaljevic et al. 2021). Briefly, the model is a set of differential equations that describes the flow of host individuals from susceptible to infectious to recovered via immunity, with the potential for serious illness to lead to hospitalization and mortality (Fig. 1). This model version does not include vaccination, although the model could be extended in the future to include vaccination or other current dynamics of COVID-19 (e.g., multiple viral strains). In the Supplementary Information, we show the full model equations, provide definitions for the state variables and parameters, and show our default parameter values used in the simulation study described herein.

**Figure 1:**
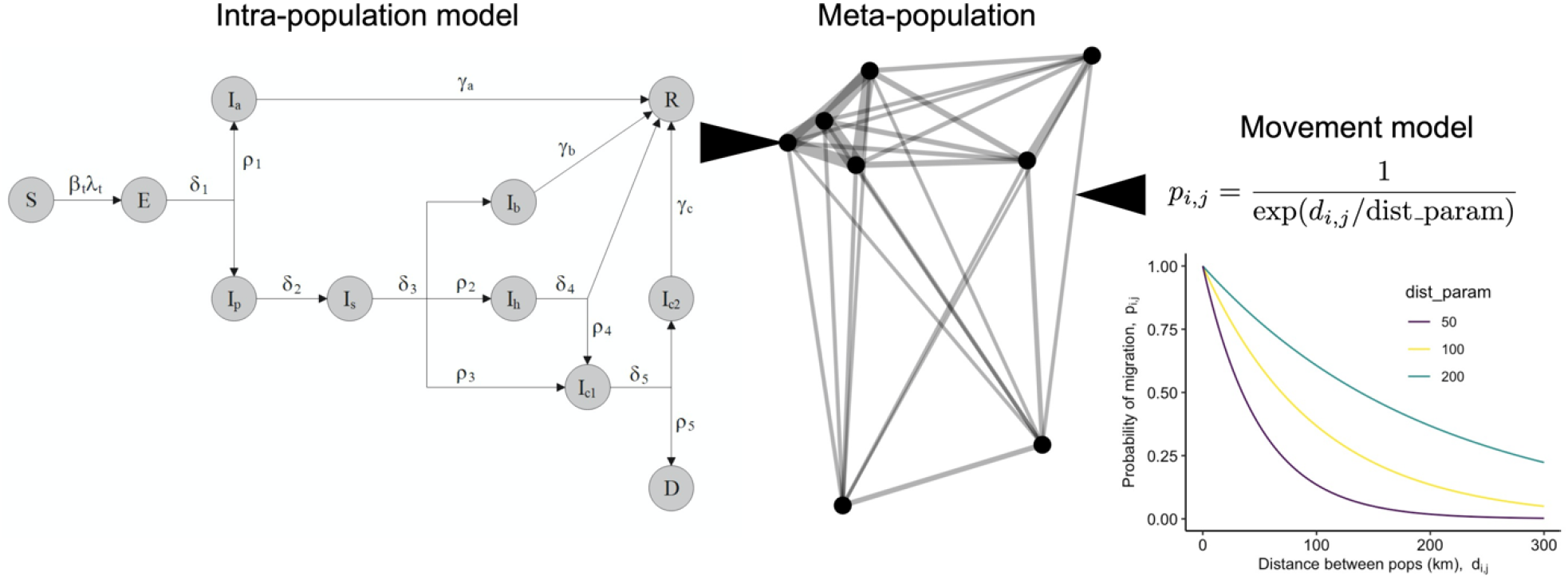
COVID-19 meta-population model schematic. The intra-population model is solved concurrently in each sub-population (node) of the meta-population. The state variables represent transition from Susceptible (S) to Exposed (E) to various Infectious classes (I) (asymptomatic (a), pre-symptomatic (p), symptomatic (s), recovering at home (b), hospitalized (h), in the ICU (c1), in the ICU step-down/recovery unit (c2)) to Recovery (R) or Death (D). Movement of susceptible and infectious host individuals can move the pathogen among sub-populations, and the probability of movement between any two populations is described by a distance-based dispersal kernel.

The model is spatially-explicit, with host movement driving spatial transmission, so we must consider the spatial characteristics of host populations in the fitting process to estimate host movement rates. The model allows host movement rates and local transmission rates to vary daily, although here we assume rates vary on a weekly basis. These time-varying rates help account for non-pharmaceutical interventions and/or behavioral changes that influence host movement between sub-populations and the probabilities of host-to-host transmission. Furthermore, the model is stochastic, meaning that, in fitting the model to data, we need to account for variability observed across stochastic realizations of the model. Upcoming sections discuss these issues in more detail.

### Spatial and temporal model dynamics

The model allows for spatial dynamics in which hosts move between discrete sub-populations. Susceptible hosts in a sub-population can therefore get exposed to the pathogen by visiting a different sub-population or by infectious hosts from different sub-populations who visit their home sub-population. We denote the per-capita movement rate by *m*, which describes how many susceptible and infectious hosts, on average, move between sub-populations per time step. This rate is not explicitly shown in the intra-population model, but we use it in a Poisson draw to determine how many individuals will move out of a given sub-population per time step. After we determine how many individuals will move from each sub-population, we use the following distance-based dispersal kernel to control the probability of movement from population *j* to population *i*.

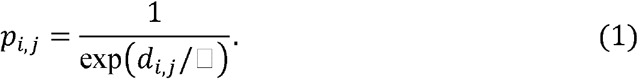

The value of *ϕ* modulates how strongly distance affects movement probabilities (e.g. the extent to which the number of roadways or transport options overcome raw separation distance), and *d*_*i,j*_ is the Euclidean distance between populations *i* and *j* (Fig. 1). We use the number of individuals that will move and these *p*_*i,j*_ values in a multinomial random draw to determine how many individuals will move between each sub-population pair per time step. In the simulations that we present below, we fix the *ϕ* (i.e., we assume that it is a known parameter), but we estimate a time-varying per-capita movement rate *m*_*t*_. Future studies could address how our fitting methods are capable of simultaneously estimating per-capita movement rate and the parameter *ϕ* in real settings.

The model also allows for a time-varying local transmission rate, *β*_*t*_. Thus, both the host movement rate *m*_*t*_ and the local transmission rate *β*_*t*_ can influence temporal patterns of COVID-19 related hospitalizations, just as in real human populations in real geographic regions. It is important to note that *β*_*t*_ and *m*_*t*_ do not vary per sub-population, although this could be the case in future studies.

Although modeling time-variation of both local transmission and inter-population movement rates has strong potential for increasing model acuity and sensitivity, it does introduce potentially confounding causal impacts on model predictions. For instance, increasing the local transmission rate and the regional movement rate may have similar effects on the observed number of new hospitalizations per time. Therefore, we must validate whether these two parameters are sufficiently identifiable when fitting the model to geo-referenced surveillance data. More broadly, our aim is to develop a generalizable new approach that can untangle causality issues like this, while simultaneously estimating time-varying rates of transmission and host movement from hospitalization data.

### Model-fitting algorithm

Our model-fitting methods rely on grid search and a sliding window approach for estimating time-varying transmission and movement rates, validated against geo-referenced data on new hospitalizations per day. We chose to focus on hospitalization data because, from our experience, they tend to be more reliable than case data, which are subject to various biases including spatially and temporally fluctuating sampling efforts. Briefly, our method relies on estimating the time-varying parameters using time slices (i.e., a sliding window) of hospitalization data, to account for the fact that changes in transmission rate or movement rate will have delayed effects on the temporal trends in hospitalization. We also rely on high-performance computing, running many (thousands of) grid searches concurrently (i.e., in an embarrassingly parallel process) to efficiently search a large parameter space (Fig. 2).

**Figure 2:**
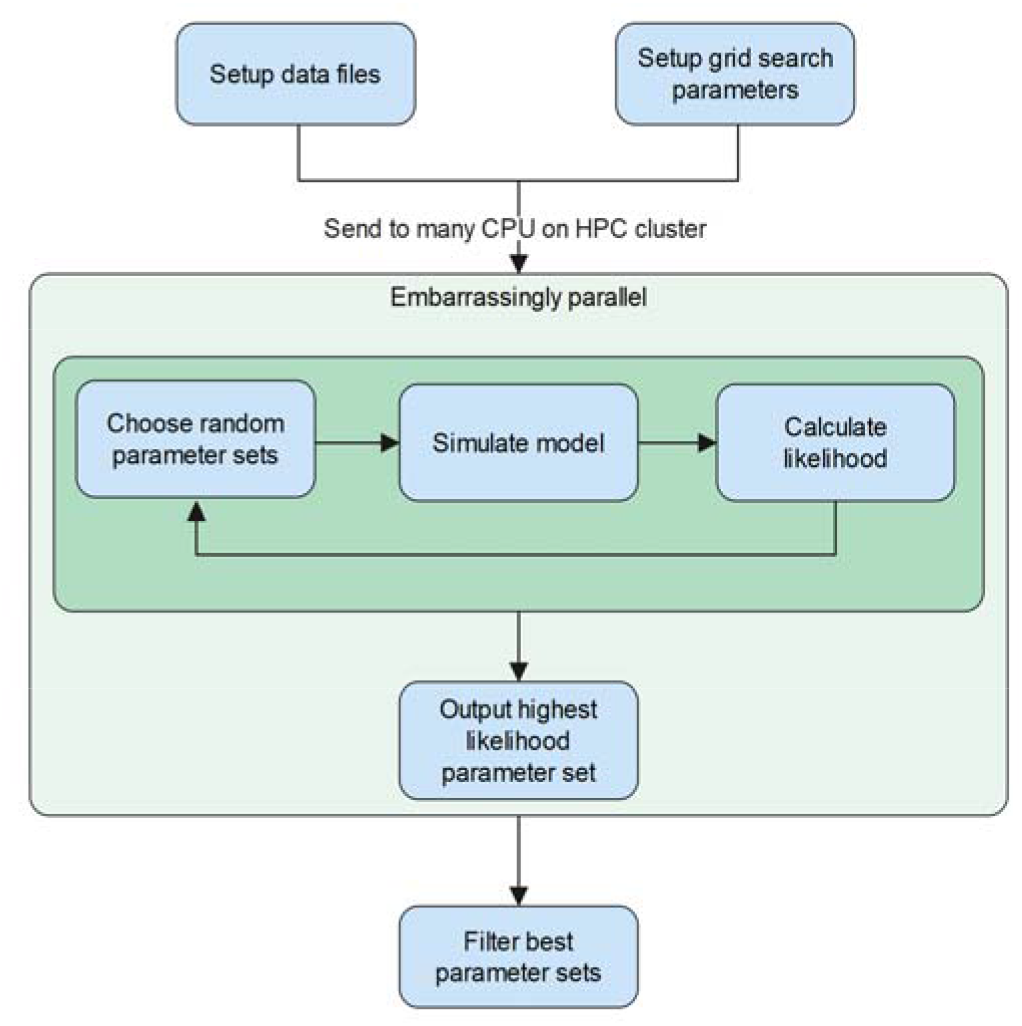
Implementation scheme of the grid search process on a high-performance computing cluster (HPC). On many CPUs simultaneously, we randomly search through the parameter space, and each CPU outputs a single best parameter set (based on likelihood calculations) that was discovered from its independent search process.

To better illustrate our sliding-window approach, suppose that we are interested in fitting *n* weeks of hospitalization data to estimate weekly-varying transmission rates *β*_*t*_, where *t* designates the week of interest. We start the grid search for the first (weekly) value *(β*_0_) based on the first three weeks of hospitalization data (Fig. 3). We chose a 21-day window-size because COVID-19 disease dynamics show an approximately three-week delay between exposure to SARS-CoV-2 and hospitalization due to COVID-19, i.e., any increase in transmission rate now may not lead to increased hospitalizations for three weeks. The random grid search tests many values of *β*_0_ across the parameter range. Each time we test a new parameter value, we re-run the model many times and compare these stochastic realizations to the hospitalization data using a likelihood method described below. We fix *β*_0_ to the best value found during that particular search, and the 3-week sliding window is then advanced by one week. We then conduct a random grid search to find the second week-specific transmission rate (*β*_1_). In this case, we would run the model spanning four weeks, fixing *β*_0_ to the best value estimated for the first week of the model simulation, and inserting the new test value of *β*_1_ for the following weeks. We continue this process, updating each weekly value of *β*_*t*_ in succession until we reach the final week of data. The full, step-wise algorithm is laid out in more detail in the Appendix.

**Figure 3:**
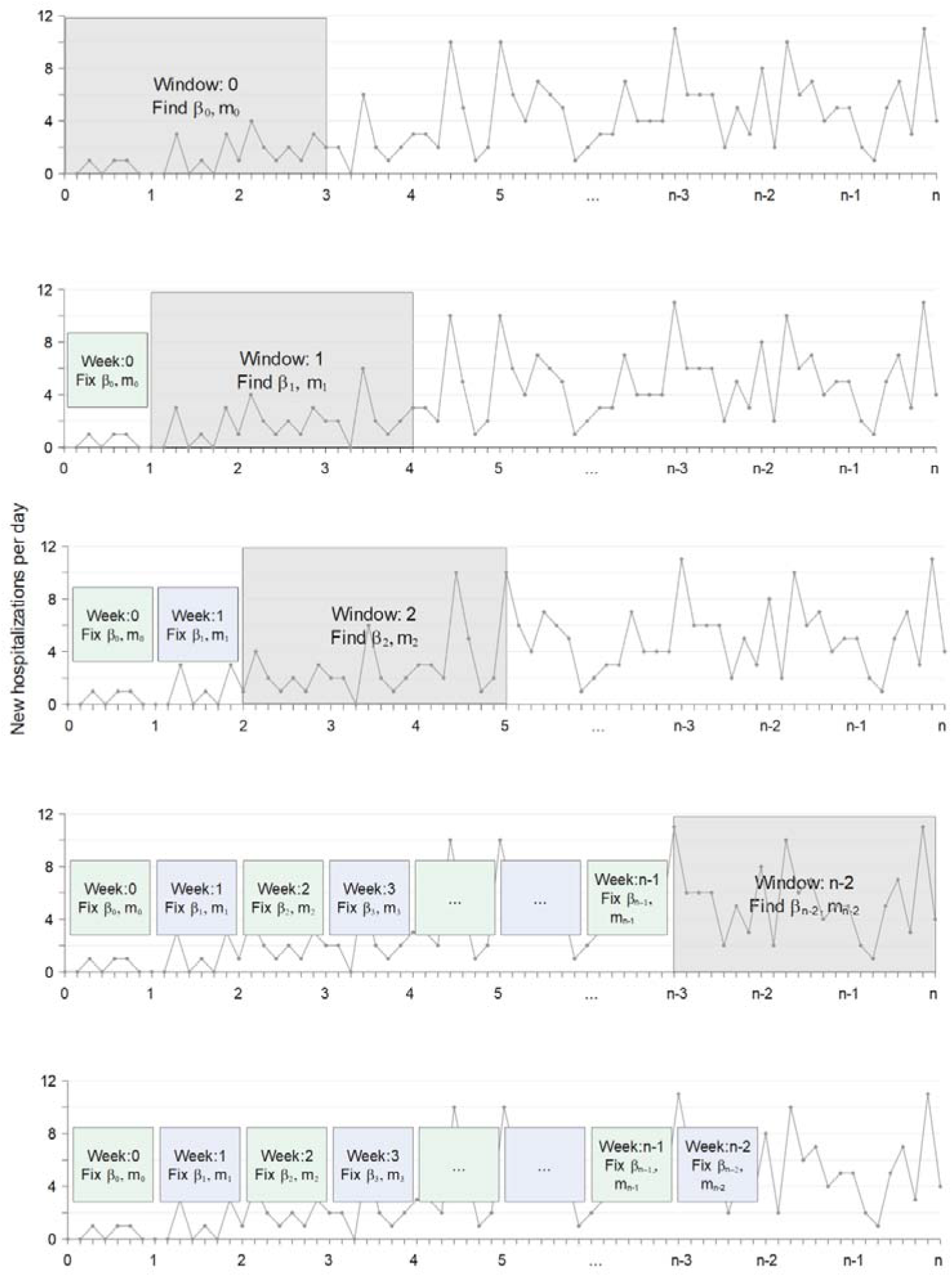
Illustration of the sliding window method with grid search to estimate the transmission rate β_t_ and the movement rate m_t_, concurrently for n −2 weeks.

In the interest of clarity, we have thus far left out a complicating detail in this example: we estimate a value of *both β*_*t*_ and *m*_*t*_ in each sliding window (Fig. 3). For each window, the process randomly chooses *β*_*t*_ or *m*_*t*_ to work on first, while the other parameter is set to its value from the previous window. Suppose *β*_*t*_ is chosen first. The process randomly searches across the range of possible *β*_*t*_, finds the best value, and then repeats a search for *m*_*t*_. Within each window, this process is repeated multiple times, choosing which parameter is searched for first, such that any confounding effects of *β*_*t*_ and *m*_*t*_ are disentangled. In this way, the model considers the additive impacts of movement and transmission rates on observed hospitalization data in each consecutive time window.

For each time window, the best parameter set (i.e., a value of *m*_*t*_ and *β*_*t*_) is chosen by calculating a likelihood score that compares the stochastic, spatially-explicit model outputs to the value of new daily hospitalizations observed in each sub-population. In this case, we compare the model to these data using a Poisson likelihood, as the hospitalizations are count data. Although we could use the negative binomial likelihood, using the Poisson was sufficient and no over-dispersion was observed in the model outcomes.

The model is stochastic, so the model is executed multiple times (i.e., for multiple realizations) every time a new parameter value is tested. For each stochastic realization, we output the number of new hospitalizations per sub-population to construct a spatially-informed likelihood that averages across the model realizations. Part of our challenge was understanding how best to structure the likelihood equation to account for data coming from distinct sub-populations. We therefore tested two separate approaches: one that tests model performance against each sub-population independently, and one that groups sub-populations based on the distance from the original epicenter sub-population (i.e., we seeded the pathogen into one epicenter sub-population, and then allowed host movement to spread the pathogen regionally between sub-populations). The reasoning for testing these two distinct likelihood approaches is rooted in how we model host movement. Because host movement is stochastic and based on pairwise distance, it is possible that the model-fitting routine would be unable to reliably match new hospitalizations per sub-population. In other words, it is more likely that the model can capture – on average – how far the pathogen may move over time, but not necessarily the exact sub-populations to which the pathogen will move per time step. More generally, the pathogen is expected to move from the epicenter towards more distant sub-populations over time, based on the dispersal kernel assumption, but this process is random.

Our first likelihood structure therefore treats each sub-population independently:

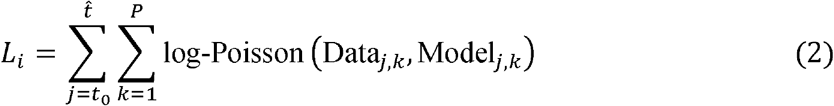

Here *L*_*i*_ is the log-likelihood for one model realization, *t*_0_ is the start time, 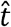 is the day upon which the simulation ends (i.e., the end of the time window), and *P* is the number of sub-populations. Thus, Data_*j,k*_ is the number of new hospitalizations recorded in sub-population *k* on day *j*. We then average the likelihood across the stochastic realizations:

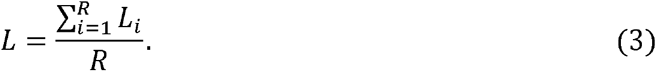

Here, *L* is the average log-likelihood for a given parameter set, and *R* is the number of model realizations that we simulate per parameter set.

The likelihood expressed in equation 2 accounts for each sub-population individually. In other words, the model keeps track of the number of daily hospitalizations per sub-population, and the likelihood calculation compares the model’s output of hospitalization per sub-population to the true value of hospitalization per sub-population from the data. Given the randomness of host movement, as described above, we tested an alternative likelihood structure in which we grouped sub-populations based on their radial distance from the known epicenter sub-population (Fig. A1). In this case, we sum the number of hospitalizations across sub-population groups:

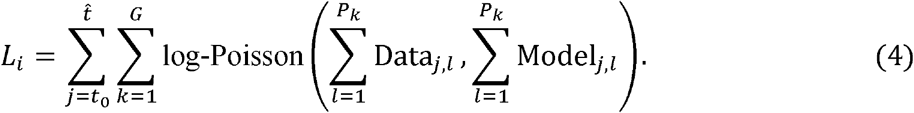

In equation 4, *G* represents the number of sub-population groupings, and *P*_*k*_ represents the number of sub-populations in group *k*, such that *l* refers to the sub-populations in group *k*. Thus, we sum the number of new hospitalizations on day across all of the sub-populations in group *k*. Here, we grouped populations based on a circular buffer of 50km (Fig. A1).

Overall, the grouped spatial likelihood structure (equation 4) provided a superior fit, so we used it for all analyses presented below. An explicit comparison between model fits when we used likelihood equation 2 (non-spatial) versus equation 4 (spatial) is included in the Appendix. Future studies could however use either likelihood structure effectively

### Simulated data and analysis

Our overall goal was to test whether we could reliably estimate both a weekly-changing transmission rate and human movement rate by fitting our spatial and stochastic model to spatially-referenced hospitalization data. To conduct these tests under an idealized scenario, we simulated hospitalization data with known model parameter values and then used our fitting routine to determine if we could recover accurate estimates of these known parameters. We simulate data from three different geographic region types: urban, suburban, and rural (Fig. 4). The urban area has sub-populations with larger numbers of individuals grouped within a smaller geographical area (population size mean (range): 205,745 (448-3,252,452); total population size: 4,732,148; spanning 8,413km^2^), whereas the rural area has sub-populations with smaller numbers of individuals scattered across a larger spatial area (population size mean (range): 2,190 (201-6,527); total population size: 72,292; spanning 9,778km^2^), with the suburban area as an intermediate between the two (population size mean (range): 63,160 (496-1,019,882); total population size: 1,136,887; spanning 6,968km^2^). The population sizes and spatial distributions of these three regions were based off spatially shuffled population data from three real counties in Arizona (Maricopa, Pima, and Apache). The sub-population locations and population sizes were originally generated from a 1km^2^ global data set on population size (Center for International Earth Science Information Network – CIESIN – Columbia 2018). These 1km^2^ values were clustered to make larger sub-populations. Then, we shuffled the latitude and longitudes of these sub-populations to randomize their locations. However, once we shuffled the locations, we used the same spatial arrangement of sub-populations for all the simulations of the disease model described below. Data on the locations and population sizes of all sub-populations can be found in the GitHub repository.

**Figure 4:**
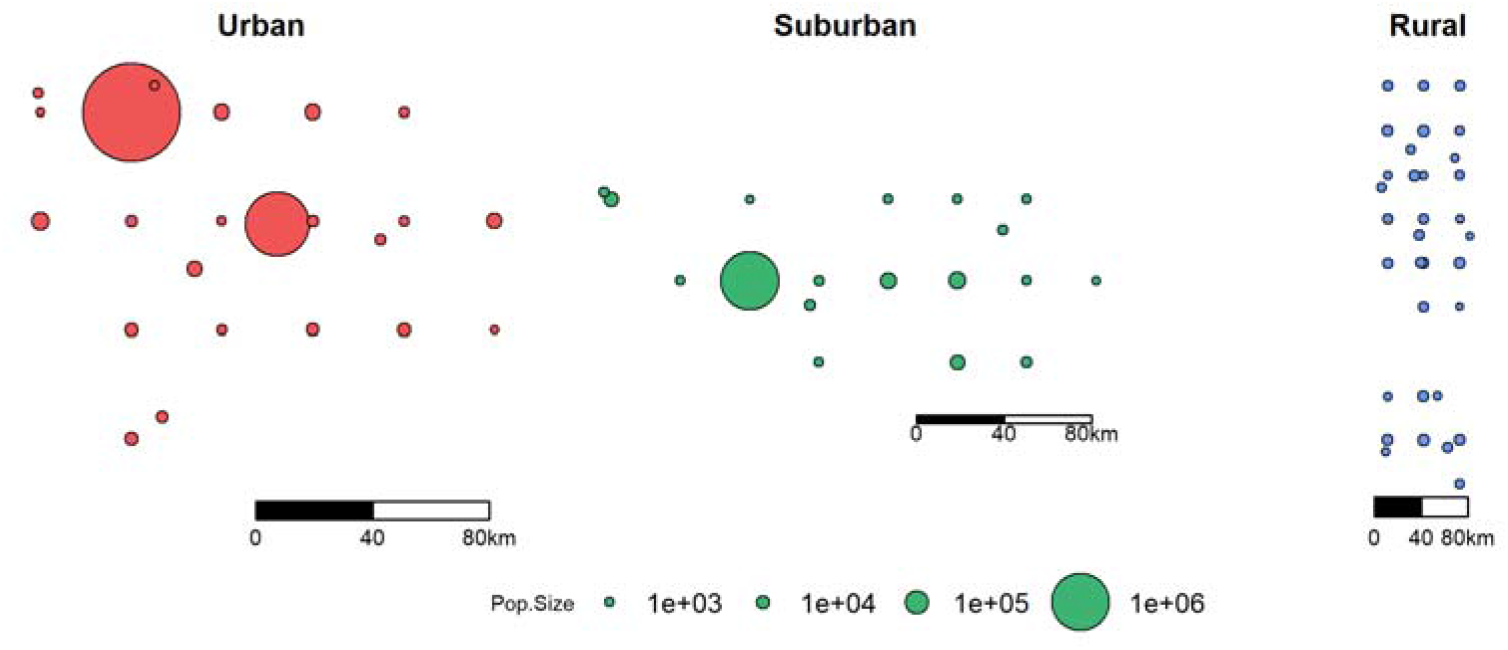
Simulated regions with different population densities and spatial structures.

We consider three situations for each of the three regions. First, we fix the weekly-varying, per-capita movement rate *m*_*t*_ to known values, while we estimate the weekly-varying transmission rate *β*_*t*_. Second, we fix *β*_*t*_ while we attempt to estimate *m*_*t*_, and third, we attempt to estimate both time-varying parameters concurrently. These three experimental conditions will help us better understand the potential for untangling the impact of the two parameter values, considering that both parameters could have similar effects on local hospitalization rates. We imposed realistic patterns of *β*_*t*_ from local COVID-19 data across Arizona, and for *m*_*t*_, we assumed low initial movement (due to, for example stay-at-home orders), which then return rather quickly to a “normal” baseline, which was a pattern seen across the USA (Google 2020; Gibbs et al. 2021). We fit the model to the simulated data from each of the three region types separately, but using the same parameter values and the same conditions in each grid search. We set the grid search range of *β*_*t*_ to [0.001,0.6] and range of *m*_*t*_ to [0.0,0.08]. We simulated a total duration of 296 days starting from 03-01-2020 for the epidemic and we used a 3-week sliding window in the fitting routine; we thus estimated parameter values for 40 weeks. We implemented the algorithm on 5000 independent CPUs on a high-performance computing cluster, with 10 grid searches and 20 realizations per week per CPU. Each CPU outputted a single best parameter set from the overall search. From this global search, we used the best 200 parameter sets (based on overall likelihood) to simulate the model and visualize model fit.

## Results

In general, our model-fitting routine does a very good job of accurately recovering weekly-varying values of the transmission rate *β*_*t*_, regardless of whether the movement rate *m*_*t*_ is known (fixed) or simultaneously estimated (Fig. 5-7). Additionally, in all scenarios, the model-fitting routine showed a high match between the model-estimated hospitalization dynamics and the actual (simulated) data. There was less precision in our method’s estimation of *m*_*t*_, regardless of whether *β*_*t*_ was fixed to known values or estimated simultaneously. While the average tendency tended to match the correct pattern of *m*_*t*_ when *β*_*t*_ was fixed, there was lower precision of *m*_*t*_ when both rates were estimated simultaneously. Interestingly, we can see that when the hospitalization numbers increase due to an increase in transmission rate (*β*)_*t*_, the fitting algorithm over-estimates *m*_*t*_ but slightly under-estimates *β*_*t*_ in those same time-periods (in the scenario where both parameters are being estimated simultaneously). This highlights and validates a concern outlined earlier, namely, the difficulty of accurately separating the effects of multiple simultaneously estimated parameters, when those parameters can have similar effects on model outcomes. In this case, movement and transmission rates can have similar effects on hospitalization patterns, and it seems that the analysis tended to favor movement over infection rate in rationalizing hospitalization patterns. Finally, and as expected, our precision of estimating transmission and movement rates was generally lower for the suburban and rural regions compared to the urban region, most likely because these regions had lower total hospitalization numbers, and we therefore had less statistical power to estimate model parameters.

**Figure 5:**
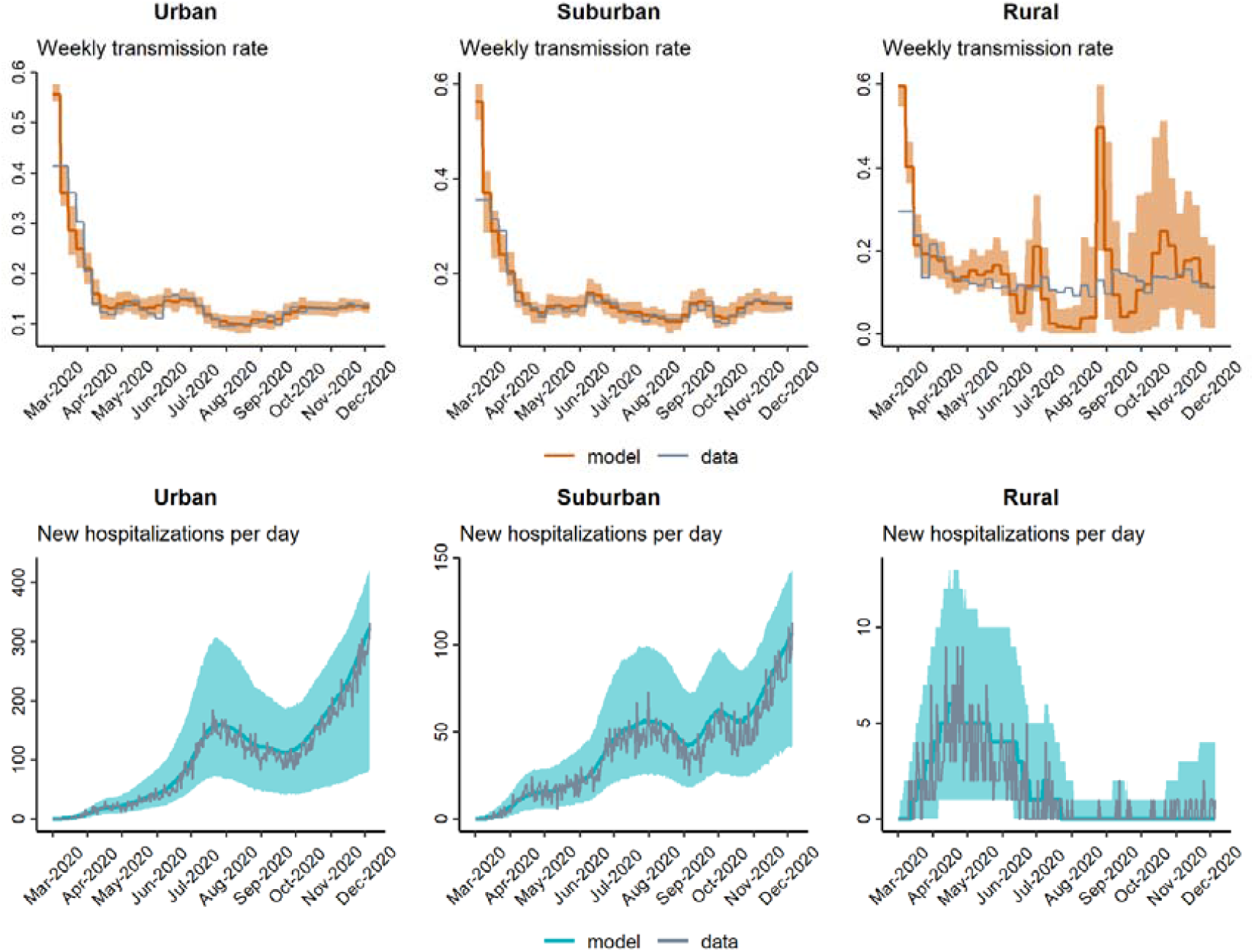
Model-fitting of weekly transmission rate β_t_ and daily hospitalizations when movement rate m_t_ is set to known values.

**Figure 6:**
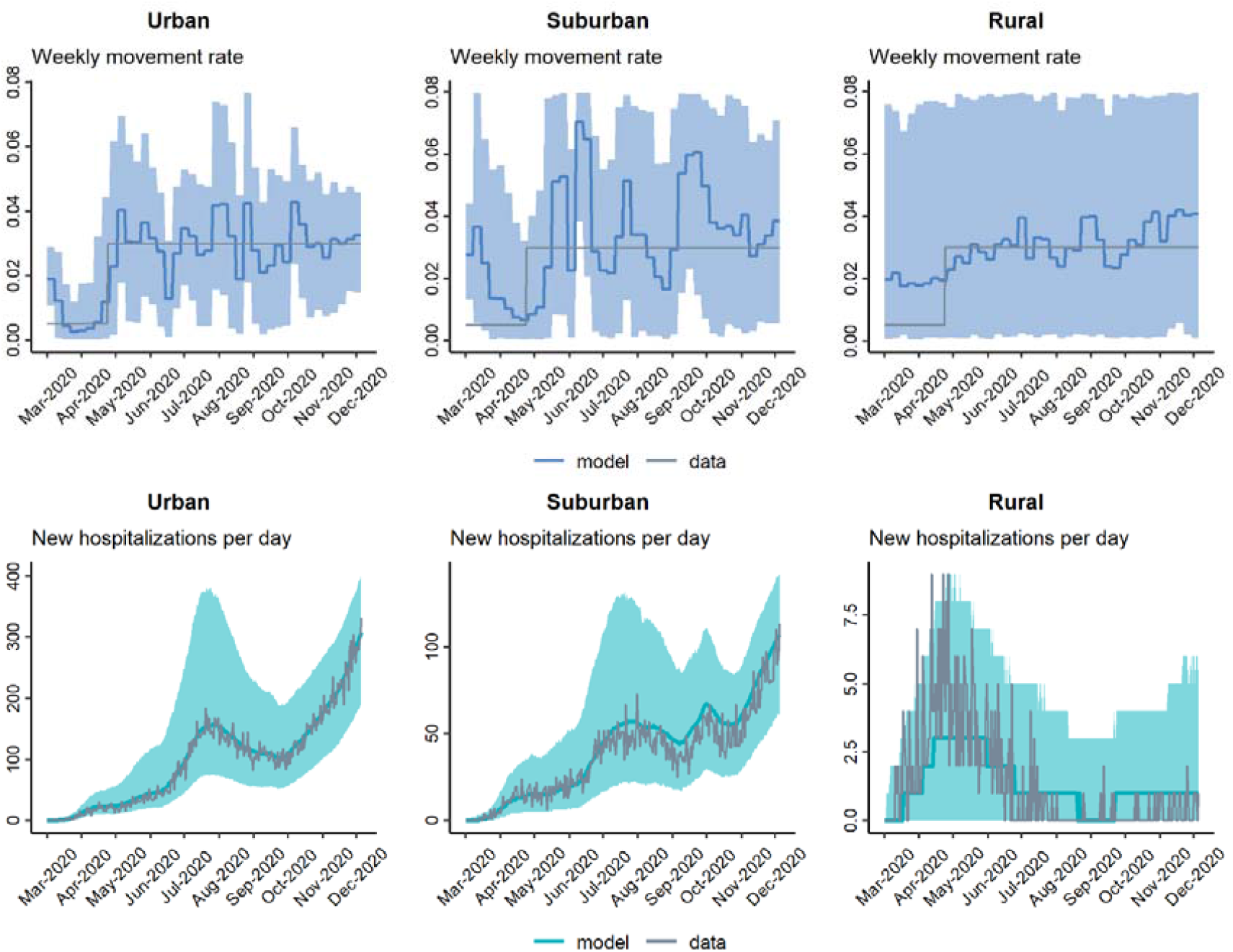
Model-fitting of weekly movement rate m_t_ and daily hospitalizations when transmission rate β_t_ is set to known values.

**Figure 7:**
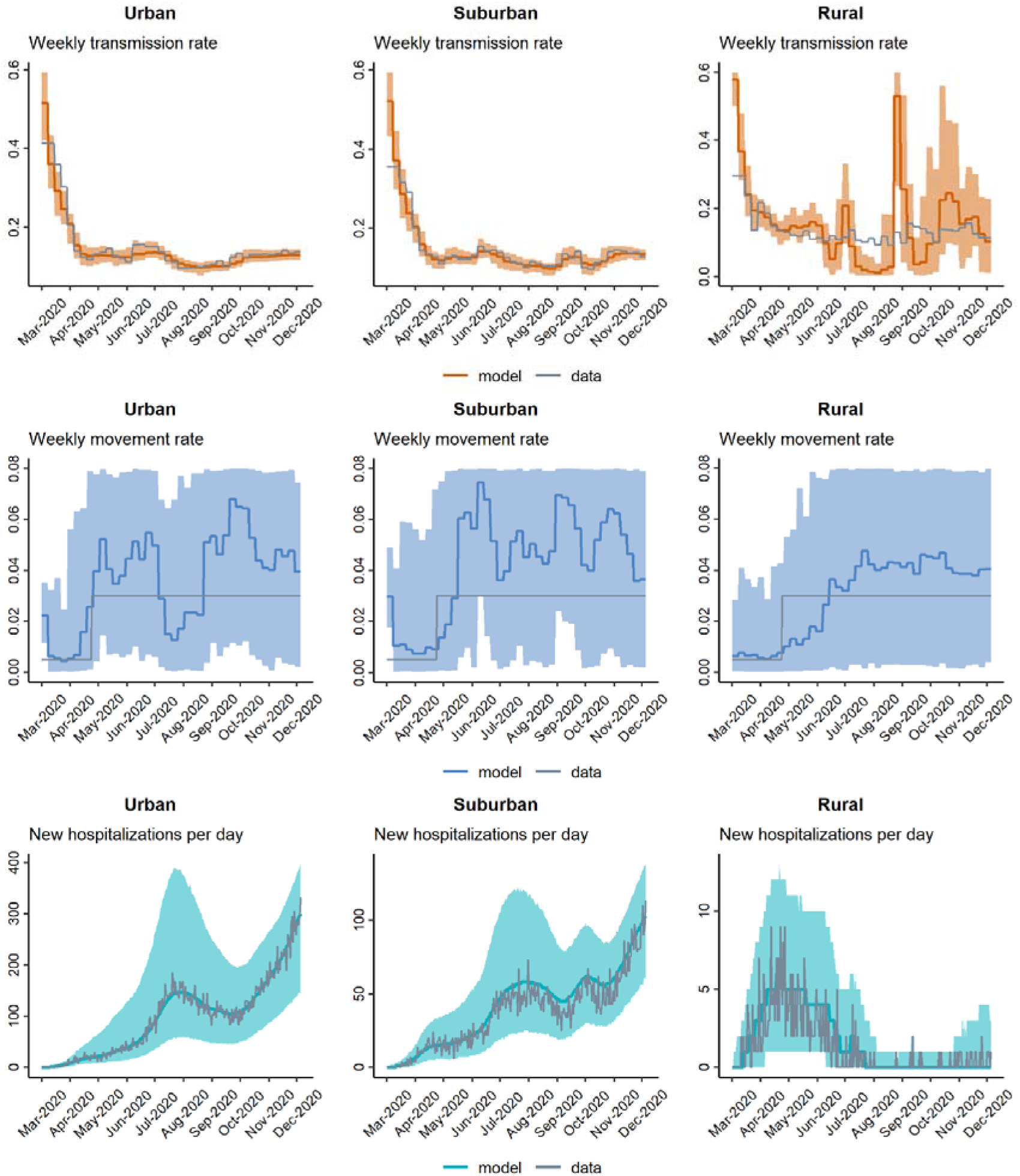
Model-fitting of weekly transmission rate β_t_ and movement rate m_t_ concurrently.

## Discussion

In this work, we explore a method to fit a spatially-explicit, stochastic model of COVID-19 to location-specific hospitalization data to estimate time-varying parameters. Specifically, we address whether the method allows us to simultaneously estimate time-varying transmission rates and time-varying movement rates. Using grid search and a sliding window technique, our method estimates weekly-varying transmission rates with high accuracy regardless of the variability of the movement rate. On the other hand, the accuracy of movement rate estimation is comparatively lower, although the fitting algorithm does capture important overall patterns in time-varying movement rates. We also show that the accuracy of model fitting varies by geographic context and population densities: the fit is most precise in high-density geographical regions (e.g., suburban to urban settings), and there is more error in low-density (e.g., rural) settings. This is likely due to lower data availability in these rural settings (e.g., lower total hospitalizations over time), reducing the method’s statistical power to estimate parameters. Our model-fitting routine offers an effective and efficient method for fitting non-linear, stochastic and spatially-explicit models to longitudinal data, which is not limited to mathematical models of COVID-19 or even epidemiology.

Grid search is a reliable parameter optimization method in low dimensional spaces and is also simple to implement in code. When executed in parallel, grid search becomes highly effective and allows more than just a point estimate. In our routine, we use a high-performance computing environment to execute many grid search processes in parallel, allowing us to quantify a likelihood profile (i.e., probability distribution) for each parameter. Moreover, in our algorithm, we added various aspects of randomness that increased the ability of the algorithm to effectively explore a broad parameter space in a time-efficient manner. Overall, our algorithm shows promise for estimating time-varying parameters, and it is easy to modify the underlying code to work with different spatial and stochastic models in epidemiology and beyond. We envision future projects that will allow us to wrap this fitting routine into a generalized parameter fitting framework, in which users could insert their model of interest, data source, and likelihood function to automate the process of parameter optimization.

Our method was not as ideal for simultaneously estimating time-varying transmission and host movement rates from a single source of data. This is a common problem in estimating model parameters that may have correlated or similar effects on the model dynamics. Fortunately, our fitting algorithm is flexible enough that in future projects we can construct a likelihood structure that includes more than one source of data. In particular, the movement and transmission rates would likely be more separable if we leveraged a movement-specific data source. For human pathogens, mobility data is becoming much more common thanks to electronic devices that include GPS. For instance, several studies have recently used mobility data from “SafeGraph” to inform epidemiological models of COVID-19 (Sen et al. 2021; Yang, Shaff, and Shaman 2021). In addition, our likelihood function did not account for spatial relations between populations except for how the populations were situated in reference to the disease epicenter. As the epidemic spreads locally and regionally, the distance from the disease epicenter becomes less relevant, as other populations may become more important epidemic hubs. Therefore, improving our likelihood function to more explicitly account for spatiotemporal auto-correlation (e.g., Shariati et al. 2020; Kim and Castro 2020) as the epidemic progresses may improve parameter identifiability. We also did not do rigorous studies to test whether the spatial arrangement of sub-populations could influence our ability to estimate transmission or movement rates. Future studies could address how different likelihood structures and the use of real mobility data could improve inference using our methods.

In emerging epidemic and pandemic situations, allowing for time-varying parameter values of epidemic models is critical for multiple reasons. If the purpose of the modeling exercise is to make short-term forecasts of disease outcomes, then accurately representing the transmission dynamics “up to present” (i.e., accurately hindcasting) is imperative to ensure that the “future” model dynamics account for temporally auto-correlated effects. Furthermore, it is important to have a reliable method to quantify the effects of specific public health interventions on model dynamics, for retroactively or even proactively understanding and modeling their impacts on epidemic patterns. Our method offers an efficient and substantially automated method to update model fits and forecasts for complex, spatially-explicit and stochastic models that should be useful in a variety of academic and applied contexts.

## Data Availability

All data produced in the present study are available upon reasonable request to the authors

## Acknowledgements

This material is based upon work supported by the National Science Foundation under Grant No. 2028629 (JRM, ED, CH) and by the Flinn Foundation under Grant No. 2305 (JRM). The funders had no role in study design, data collection and analysis, decision to publish, or preparation of the manuscript.

## Supporting Information List of Legends

**Table A1:** *COVID-19 model state variables*

**Table A2:** *COVID-19 model parameter descriptions*

**Table A3:** *Algorithm for grid search with sliding window technique*

**Figure A1:** *Schematic that shows how populations were grouped by a 50km buffer around the epicenter sub-population (shown in red), used to calculate the group-based likelihood (equation 4). In this case, hospitalization data from sub-populations in the same group (shown as letters) are added together*.

**Figure A2:** *Estimate of β*_*t*_ *when m*_*t*_ *is known, comparing our two likelihood methods. Note that, although subtle, more error in the estimate of β*_*t*_ *can be detected when populations are not grouped, particularly in the rural scenario*.

**Figure A3:** *Estimate m*_*t*_ *when β*_*t*_ *is known, comparing the two likelihood methods. Note that effects of the likelihood method are not strong in this case*.

**Figure A4:** *Estimates of both β*_*t*_ *and m*_*t*_, *comparing the two likelihood methods. By grouping the populations, there is less variation in the parameter estimates, which leads to less variation in how the model fits to the data*.

