## Supporting Information for "A Sliding Window Approach to Optimize the Time-varying Parameters of a Spatially-explicit and Stochastic Model of COVID-19"

Saikanth Ratnavale *et al.*

The following set of ordinary differential equations describes the dynamics within each sub-population of the model:

$$\begin{matrix} \frac{dS}{dt} & =-\beta_{t}\lambda_{t}S-m_{t}S \\ \frac{dE}{dt} & =\beta_{t}\lambda_{t}S-\delta_{1}E \\ \frac{dI_{a}}{dt} & =\delta_{1}\rho_{1}E-\gamma_{a}I_{a}-m_{t}I_{a} \\ \frac{dI_{p}}{dt} & =\delta_{1}\left( 1-\rho_{1} \right)E-\delta_{2}I_{p}-m_{t}I_{p} \\ \frac{dI_{s}}{dt} & =\delta_{2}I_{p}-\delta_{3}I_{s}-m_{t}I_{s} \\ \frac{dI_{b}}{dt} & =\delta_{3}\left( 1-\rho_{2}-\rho_{3} \right)I_{s}-\gamma_{b}I_{b} \\ \frac{dI_{h}}{dt} & =\delta_{3}\rho_{2}I_{s}-\delta_{4}I_{h} \\ \frac{dI_{c1}}{dt} & =\delta_{3}\rho_{3}I_{s}+\delta_{4}\rho_{4}I_{h}-\delta_{5}I_{c1} \\ \frac{dI_{c2}}{dt} & =\delta_{5}\left( 1-\rho_{5} \right)I_{c1}-\gamma_{c}I_{c2} \\ \frac{dD}{dt} & =\delta_{5}\rho_{5}I_{c1} \\ \frac{dR}{dt} & =\gamma_{a}I_{a}+\gamma_{b}I_{b}+\gamma_{c}I_{c2}+\delta_{4}\left( 1-\rho_{4} \right)I_{h} \end{matrix}$$

and $\lambda_{t}$ represents a component of the force of infection, given by

$$\lambda_{t}=\frac{\omega_{1}I_{a}+I_{p}+I_{s}+I_{b}+\omega_{2}\left( I_{h}+I_{c1}+I_{c2} \right)}{N-D}.$$

**Table A1:** COVID-19 model state variables

| **State Variable** | **Description** |
| --- | --- |
| $S$ | Number of susceptible individuals |
| $E$ | Number of exposed individuals |
| $I_{a}$ | Number of asymptomatic individuals |
| $I_{p}$ | Number of pre-symptomatic individuals |
| $I_{s}$ | Number of mildly symptomatic individuals |
| $I_{b}$ | Number of mildly symptomatic individuals on bed rest at home |
| $I_{h}$ | Number of hospitalized individuals |
| $I_{c1}$ | Number of individuals in the ICU |
| $I_{c2}$ | Number of individuals in the recovery (step-down) ICU |
| $D$ | Number of deceased individuals |
| $R$ | Number of recovered individuals |
| $N$ | Total number of individuals in the population |

**Table A2:** COVID-19 model parameter descriptions

| **Parameter** | **Description** | **Value** |
| --- | --- | --- |
| $\beta_{t}$ | Time-varying transmission rate | Variable |
| $m_{t}$ | Time-varying movement rate | Variable |
| $\omega_{1}$ | Proportion reduction in transmission for asymptomatic folks | 0.55 |
| $\omega_{2}$ | Proportion reduction in transmission for hospitalized folks | 0.05 |
| $\delta_{1}$ | Transition rate: exposed to pre-symptomatic | 1/3.0 |
| $\delta_{2}$ | Transition rate: pre-symptomatic to symptomatic | 1/2.0 |
| $\delta_{3}$ | Transition rate: symptomatic to home or regular hospital bed or ICU | 1/6.0 |
| $\delta_{4}$ | Transition rate: regular hospital bed to home or ICU | 1/7.0 |
| $\delta_{5}$ | Transition rate: ICU to step-down ICU or decease | 1/8.0 |
| $\gamma_{a}$ | Recovery rate: asymptomatic | 1/6.0 |
| $\gamma_{b}$ | Recovery rate: home bed | 1/3.0 |
| $\gamma_{c}$ | Recovery rate: step-down ICU | 1/4.0 |
| $\rho_{1}$ | Fraction of exposed that transition to asymptomatic | 0.28 |
| $\rho_{2}$ | Fraction of symptomatic that transition to hospital bed | 0.20 |
| $\rho_{3}$ | Fraction of symptomatic that transition directly to ICU bed | 0.015 |
| $\rho_{4}$ | Fraction of hospitalized that transition to ICU | 0.28 |
| $\rho_{5}$ | Fraction of patients in ICU that die of disease | 0.60 |
| $\phi$ | Constant controlling the dispersal kernel shape | 25.0 |
| $\sigma$ | Stochasticity standard deviation for transmission rate (stoch_sd) | 0.05 |

**Table A3:** Algorithm for grid search with sliding window technique

**Algorithm**

1. Read raw data, decide number of days to consider.
2. Set parameter min/max, window_size and compute n_windows.
3. Set win_count = 0, param_temp and n_searches.
4. Compute temporary maximum likelihood: temp_max, initialize best parameter set.
5. Set maximum likelihood: max_lhood = temp_max.
6. Set search_ID = 0.
7. Compute number of steps and step size.
8. Set index = starter.
9. Set param = min.
10. Compute temp_max.
11. Update best parameter set for search_ID if temp_max >= max_lhood.
12. Stop if param = max, or set param = param + step size and return to Step 10.
13. Reset param_temp to current best parameter set.
14. Stop, or set index = index + 1 and return to Step 9.
15. Stop if search_ID = n_searches, or set search_ID = search_ID + 1 and return to Step 7.
16. Save the best parameter set for the current window.
17. Stop if win_count = n_windows, or set win_count = win_count + 1 and return to Step 4.
18. Calculate the full likelihood for all windows considered.

Above, we denote n_windows the number of windows for which we calculate the parameter sets. This can be obtained by, first calculating the number of weeks (n_weeks) from the raw data and then by n_windows = n_weeks - window_size + 1. Suppose that, we have $n$ number of weeks with sliding window size of 3 weeks, we find $n-2$ windows or weekly parameter sets (see Figure 3). Any remaining days beyond the final window will be included to that window, so the final window size may vary depending on the total number of days. By n_searches, we denote the number of grid searches carried out for one window. For each window, we randomly choose the starting parameter with index (starter) and optimize all the parameters, one at a time.
**Supporting Figures**

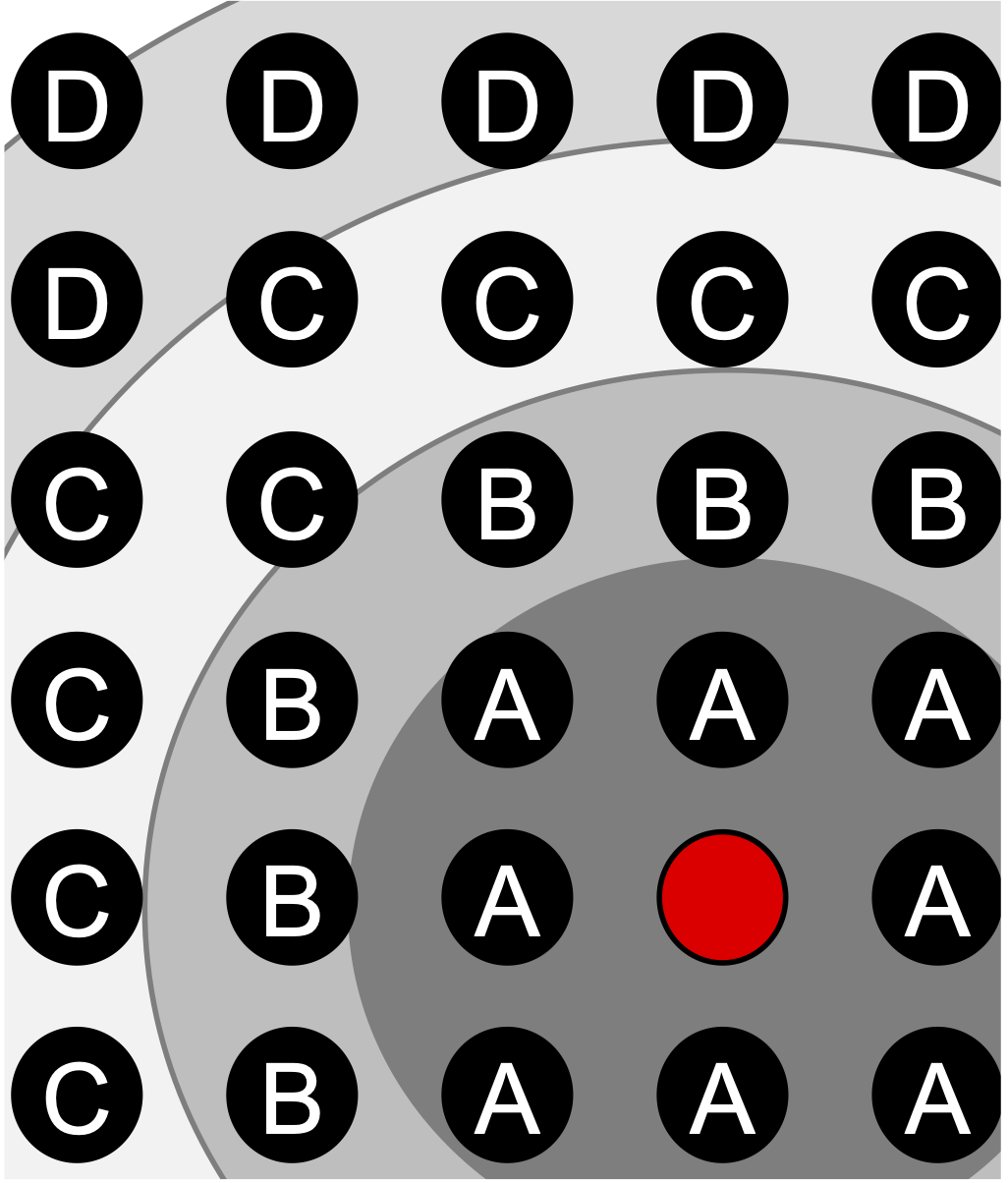

**Figure A1:** Schematic that shows how populations were grouped by a 50km buffer around the epicenter sub-population (shown in red), used to calculate the group-based likelihood (equation 4). In this case, hospitalization data from sub-populations in the same group (shown as letters) are added together.

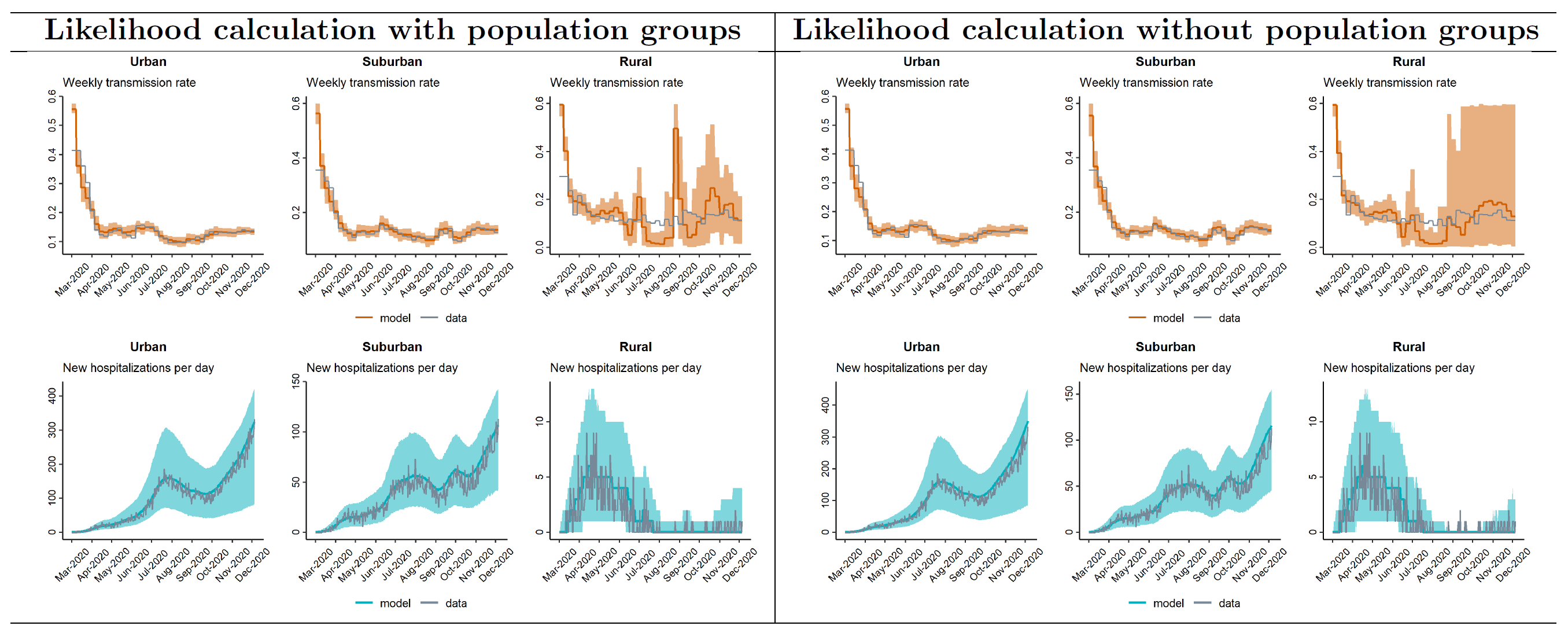

**Figure A2:** Estimate of $\beta_{t}$ when $m_{t}$ is known, comparing our two likelihood methods. Note that, although subtle, more error in the estimate of $\beta_{t}$ can be detected when populations are not grouped, particularly in the rural scenario.

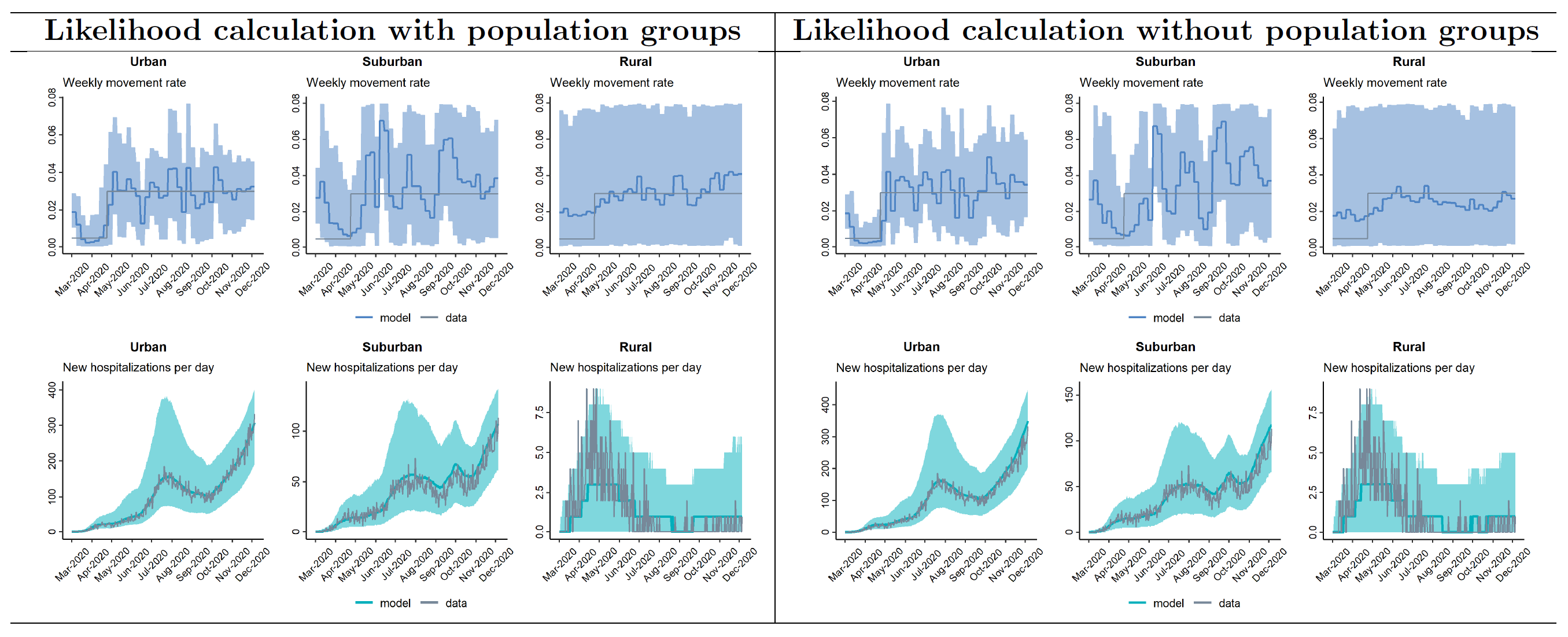

**Figure A3:** Estimate $m_{t}$ when $\beta_{t}$ is known, comparing the two likelihood methods. Note that effects of the likelihood method are not strong in this case.

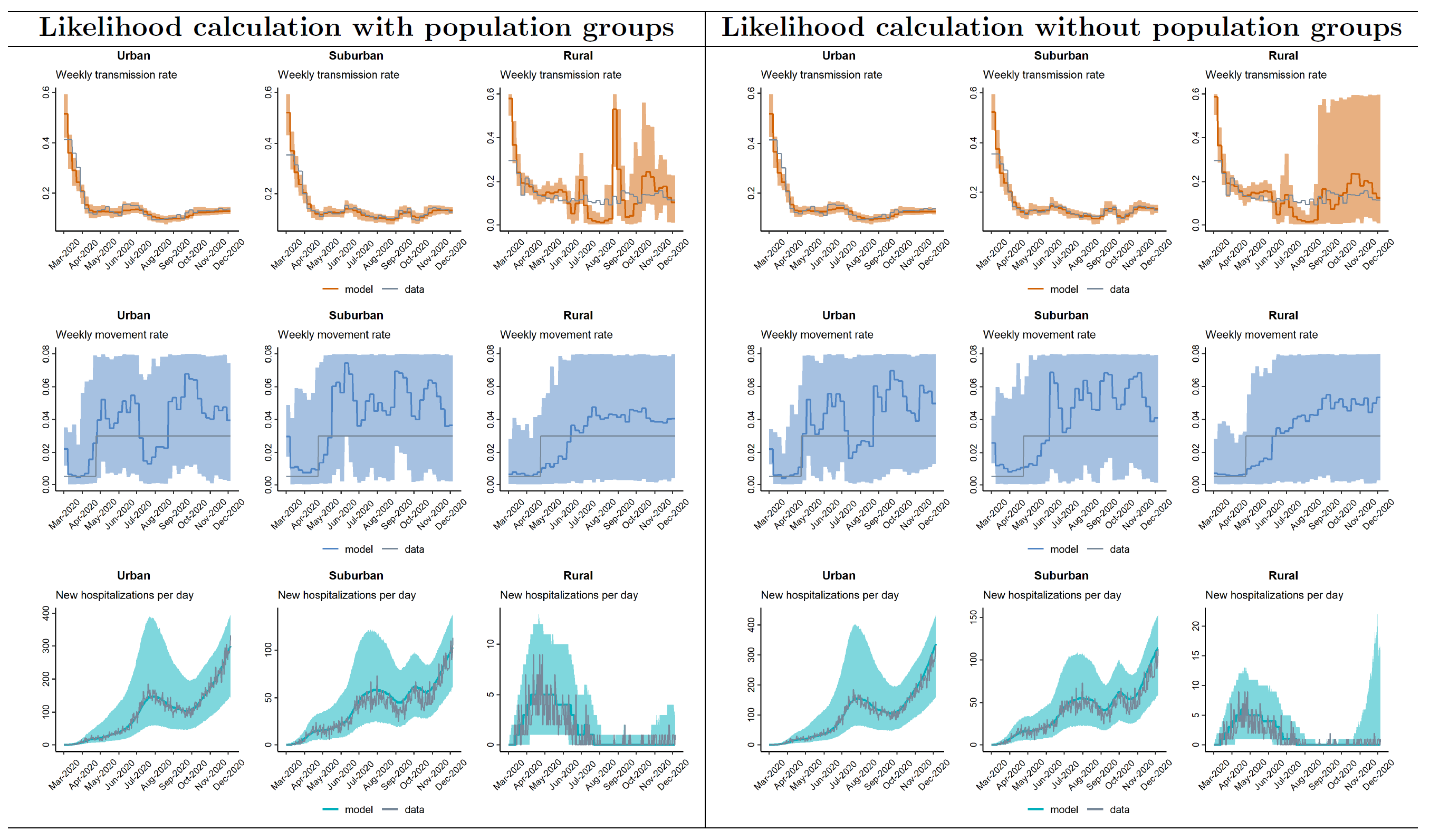

**Figure A4:** Estimates of both $\beta_{t}$ and $m_{t}$, comparing the two likelihood methods. By grouping the populations, there is less variation in the parameter estimates, which leads to less variation in how the model fits to the data.
